# Efficacy and Safety of Sitafloxacin-Containing Regimen for Improved Treatment of Nontuberculous Mycobacterial Lung Disease: A Multi-Center Retrospective Study

**DOI:** 10.64898/2026.01.13.26343918

**Authors:** Yiru Zhang, Hongxin Fu, Chenxia Zhang, Bihan Xu, Kefan Bi, Meifang Yang, Haiying Yu, Yongtao Li, Jing Guo, Wei Wu, Kaijin Xu, Ying Zhang

**Author notes:** Yiru Zhang and Hongxin Fu contributed equally to this work.

## Abstract

**Objectives:** To evaluate the efficacy and safety of sitafloxacin-containing regimens versus non-sitafloxacin therapy in patients with nontuberculous mycobacterial (NTM) pulmonary disease, focusing on sputum/BALF conversion rate, time of sputum/BALF culture conversion and radiographic improvement.

**Methods:** This retrospective cohort study analyzed 149 adults (76 control group vs. 73 sitafloxacin group) with NTM pulmonary disease treated between 2021 to 2024. Inclusion criteria: (1) Sitafloxacin group: ≥ 3 months of sitafloxacin-based therapy; (2) Both groups: Confirmed diagnosis of NTM pulmonary disease and age ≥ 18 years old. Exclusion criteria: extrapulmonary/disseminated NTM, HIV, active tuberculosis, or incomplete clinical data. Primary endpoint: culture conversion rate and time to culture conversion. Secondary endpoints: radiographic improvement and adverse events (AEs).

**Results:** The sitafloxacin group demonstrated significantly higher conversion rate (53.8% vs. 22.1%, *P* ˂ 0.001) and faster culture conversion than the control group without sitafloxacin (median 195 vs. 292 days, *P* ˂ 0.001). Radiographic improvement was more frequent with the sitafloxacin group (54.5% vs. 36.1%, *P* = 0.046). Compared to the control group, the sitafloxacin group exhibited no significant adverse events.

**Conclusions:** Sitafloxacin-based regimens accelerate microbiological clearance and promote radiographic healing in NTM pulmonary disease with good safety, positioning it as a viable drug for improved treatment of NTM infections.

## Introduction

Nontuberculous mycobacterial (NTM) pulmonary diseases, particularly caused by *Mycobacterium avium* complex (MAC) and *Mycobacterium abscessus* complex (MABC), have emerged as a growing global health challenge, with rising incidence rates linked to aging populations, especially in individuals with chronic lung disease.^1–3^ Current standard NTM treatment involves macrolide-based multidrug regimens—azithromycin or clarithromycin plus ethambutol and rifampin for MAC, and macrolides with amikacin and beta-lactams for *M. abscessus* with treatment duration lasting for 12 months post-culture conversion.^4–7^ However, while sputum culture conversion rates could reach approximately 67-84% in drug-susceptible cases for MAC and the overall sputum culture conversion in patients with MABC (34%-41% for *M. abscessus* subsp. *abscessus* and 54-69.8% for *M. abscessus* subsp. *massiliense*), treatment success remains poor in macrolide-resistant disease, with sputum culture conversion rates falling to as low as 5-36%.^7–12^ Compounding this issue, prolonged treatment durations (often > 12 months) and high rates of adverse drug reactions (ADRs) further limit therapeutic adherence and outcomes.^7,13,14^

Fluoroquinolones, particularly moxifloxacin, have been explored as alternative agents due to their potent bactericidal activity and favorable lung tissue penetration.^15–17^ Sitafloxacin, a fourth-generation fluoroquinolone, exhibits enhanced in vitro potency against NTM isolates, including macrolide-resistant strains, by dual-targeting DNA gyrase and topoisomerase IV.^18,19^ In vitro susceptibility testing has demonstrated that sitafloxacin exhibits superior activity compared to moxifloxacin, with minimum inhibitory concentration (MIC) 1-2-fold lower for MAC reference strains and 4-8-fold lower for MABC reference strains.^20^ Although preliminary evidence from a small-scale Japanese single-arm study suggests clinical efficacy of sitafloxacin in a subset of patients with refractory MAC pulmonary disease, real-world evidence regarding its effectiveness and safety for NTM pulmonary disease management remains limited.

To address this gap, we conducted a comparative cohort study evaluating the impact of sitafloxacin-containing regimens on the rate of culture conversion, time to culture conversion, radiographic improvement, and tolerability in a larger number of patients with NTM pulmonary diseases.

## Methods

### Study design and population

This multi-center, retrospective cohort study was conducted at the First Affiliated Hospital of Zhejiang University School of Medicine, Hangzhou Red Cross Hospital, Huzhou Central Hospital, Zhoushan Hospital, Wenzhou Central Hospital, Hwa Mei Hospital to evaluate the efficacy and safety of sitafloxacin-containing regimens in patients with NTM pulmonary disease. Between January 2021 and June 2024, electronic medical records were systematically reviewed to identify eligible patients diagnosed with NTM pulmonary disease according to the 2020 ATS/ERS diagnostic criteria,^7^ which require microbiological confirmation (≥ 2 positive sputum cultures for the same NTM species within 12 months or ≥ 1 positive culture from bronchoscopic sampling/lung biopsy) alongside clinical and radiographic evidence of disease (e.g., cough, hemoptysis, and characteristic CT findings such as bronchiectasis or cavitary lesions).

A total of 207 patients were assessed for eligibility (**Figure 1**), of whom 51 were excluded due to age under 18 years, pregnancy or lactation, co-infection with tuberculosis, or HIV infection. Consequently, 156 patients were enrolled and further screened (Figure 1), among whom 80 had received sitafloxacin-based therapy. After rigorous evaluation, seven patients were excluded: three due to extrapulmonary NTM involvement (bone, n = 1; synovial tissue, n = 1; pleural effusion after prosthesis removal, n = 1), one with disseminated infection, and three with sitafloxacin treatment durations of less than 3 months. Consequently, 73 patients were included in the sitafloxacin group. The control cohort comprised 76 patients who received guideline-based non-sitafloxacin regimens during the same period, matched for age, sex, NTM species, and disease severity. Inclusion criteria mandated a confirmed diagnosis of NTM pulmonary disease, age ≥ 18 years old, and a minimum of 3 months of continuous sitafloxacin therapy for patients in the treatment group. Exclusion criteria encompassed extrapulmonary/disseminated NTM infections, HIV infection (CD4^+^ count < 200 cells/μL), active tuberculosis, and incomplete treatment records.

**Figure 1.**
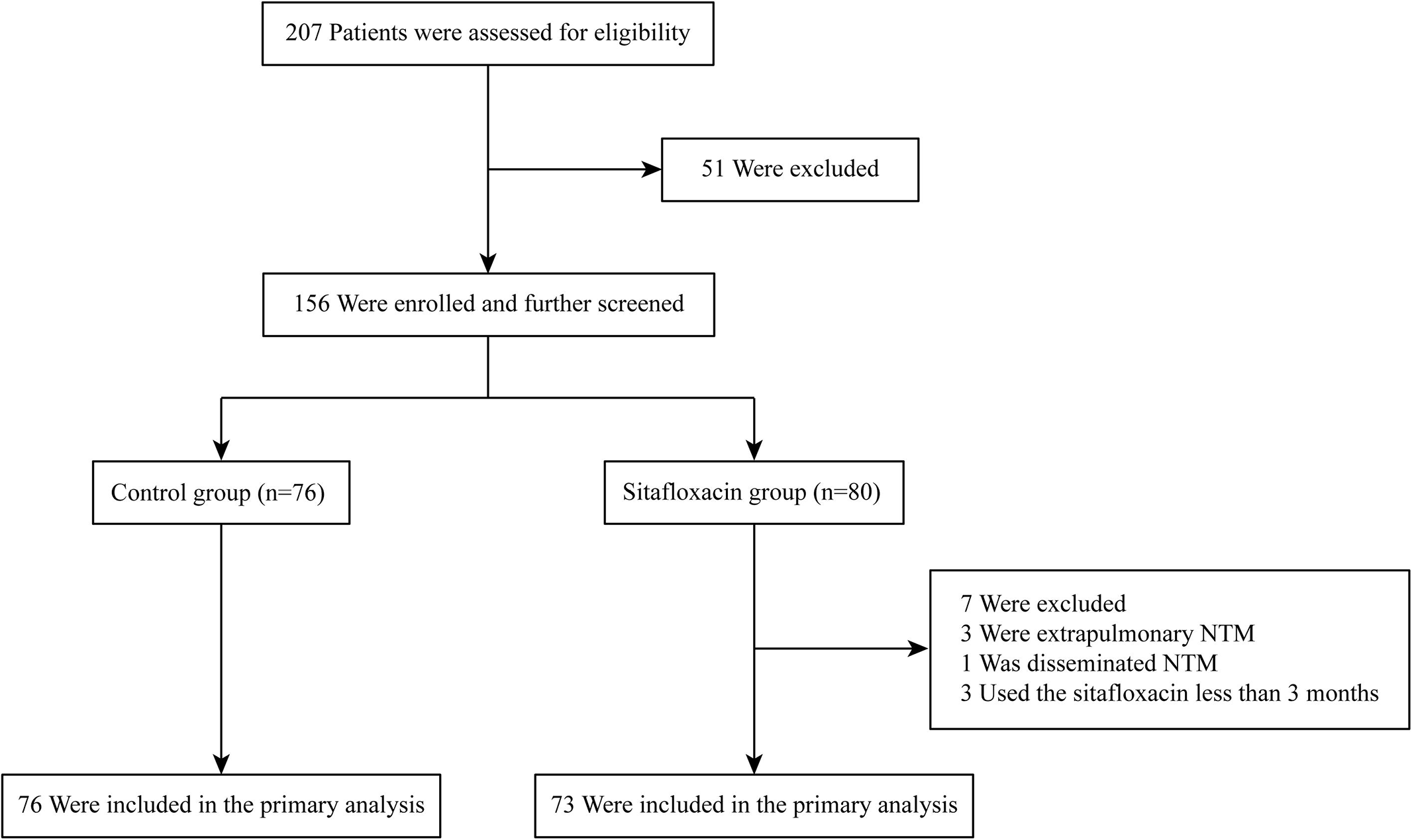
Study flowchart of patient enrollment and allocation. The research included all enrolled participants except those who failed to meet eligibility criteria. NTM, nontuberculous mycobacteria

This study was reviewed and approved by the Ethics Committee of the First Affiliated Hospital, Zhejiang University School of Medicine (Approval number: 2025B-IIT Ethics Approval No. 0666), with waived informed consent due to the retrospective use of anonymized clinical data. All procedures were performed in accordance with the ethical standards of the institutional and/or national research committee and with the 1964 Helsinki Declaration and its later amendments.

### Data collection and evaluation of treatment and safety

Nine trained researchers independently extracted data using a standardized electronic case report form, with discrepancies resolved through consensus review. Demographic variables included age, sex, smoking history (pack-years), and alcohol history. Comorbidities were documented, including preexisting pulmonary conditions (e.g., bronchiectasis, chronic obstructive pulmonary disease [COPD] and prior tuberculosis), systemic diseases (cancers and immunologic disorders), and immunosuppressive states (solid organ transplantation). Microbiological data encompassed species identification via mycobacterial DNA detection, next-generation sequencing (NGS) of BALF, acid-fast bacilli (AFB) smear, and culture of sputum/BALF, supplemented by Xpert MTB/RIF to exclude *M. tuberculosis.* Treatment-related variables included antimicrobial regimens, sitafloxacin dosing (50-200 mg/day), duration of therapy, and rationale for initiation/discontinuation (e.g., intolerance, clinical failure).

In both cohorts, outcome measures focused on sputum/BALF culture conversion, defined as three consecutive negative cultures in sputum or one negative cultures in BALF ≥ 4 weeks apart, with conversion dates documented to calculate time-to-event. The median duration of therapy before the usage of sitafloxacin treatment was 265.5 days (IQR 86.5-591.5). To ensure comparability between groups, the culture conversion rate, culture positivity and smear positivity were assessed at the median time to culture conversion in the sitafloxacin group—thereby evaluating whether patients in the control group achieved culture conversion within the same time frame. Radiologic outcomes were assessed by two independent doctors blinded to treatment allocation using pre- and post-treatment (with or without sitafloxacin) CT scans and categorized as “improved”, “unchanged”, or “worsened”. Adverse events (AEs) were monitored and categorized into systemic categories: gastrointestinal reactions (nausea, vomiting, and diarrhea), hepatic or renal dysfunction (elevated ALT/AST or serum creatinine), hematologic abnormalities (anemia, leukopenia, and thrombocytopenia), arthralgia, and rash. All AEs were systematically documented from electronic medical records, including patient self-reports during follow-up visits and clinician-observed events.

### Statistical analysis

Continuous variables were expressed as mean ± standard deviation (SD), median (interquartile range, IQR) or median (95% CI). Categorical variables were summarized as frequencies (%) and compared using chi-square or Fisher’s exact tests. The primary outcome, time of culture conversion, was evaluated by the Log-rank test. All the statistical analyses, including descriptive statistics and hypothesis testing, were performed using SPSS software (version 25; IBM Corp., Armonk, NY, USA). A two-tailed p-value < 0.05 was considered statistically significant.

## Results

### Patient characteristics

A total of 149 patients diagnosed with NTM lung disease were included in the study, comprising 76 patients in the control group and 73 in the sitafloxacin group (**Table 1**). The median age was comparable between the two groups (63.5 years [IQR 57-72.75] vs. 65 years [IQR 56-69], *P* = 0.30). The proportion of female patients was higher in both groups, with no significant difference in gender distribution (*P* = 0.70). There were no significant differences between groups in the number of smokers (5/45 vs. 6/45, *P* = 0.78), years of smoking (36 ± 13.4 vs. 37.5 ± 9.6, *P* = 0.86), average number of cigarettes smoked per day (12.6 ± 10.2 vs. 5.0 ± 10.0, *P* = 0.27), or years of quitting smoking (9.8 ± 7.5 vs. 3.5 ± 3.1, *P* = 0.18). Similarly, the proportion of patients with a history of alcohol consumption was low and not significantly different between the groups (*P* = 1.00). No significant differences were observed between groups in terms of comorbidities or past medical history, including old pulmonary tuberculosis (23.7% vs. 17.8%, *P* = 0.38), bronchiectasis (61.8% vs. 48.0%, *P* = 0.09), COPD (14.5% vs. 17.8%, *P* = 0.58), fungal pneumonia (22.4% vs. 28.8%, *P* = 0.37), cancers (15.8% vs. 11.0%, *P* = 0.39), and immune-related diseases (7.9% vs. 13.7%, *P* = 0.25). Only one patient in the sitafloxacin group had undergone lung transplantation.

**Table 1.**
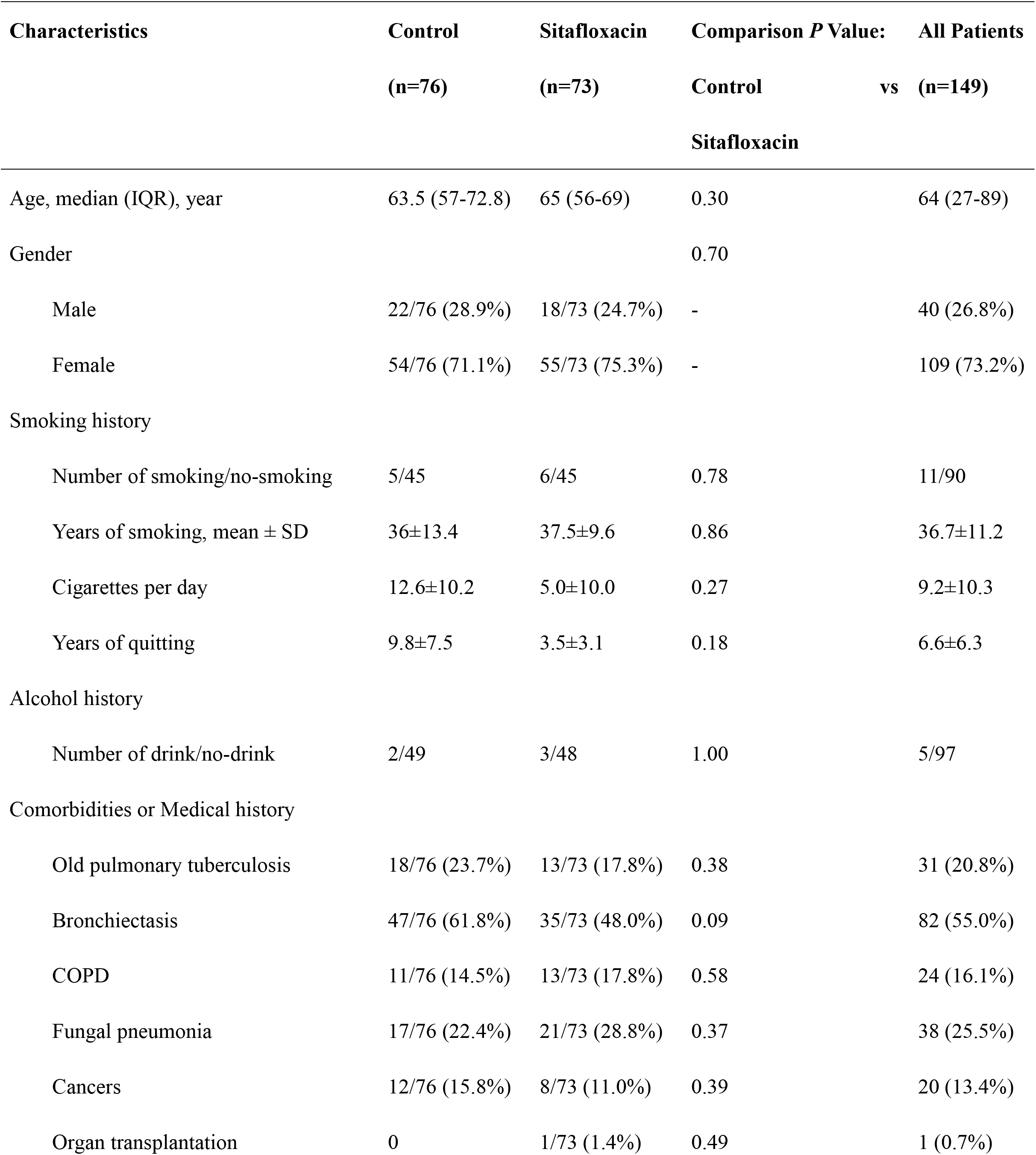
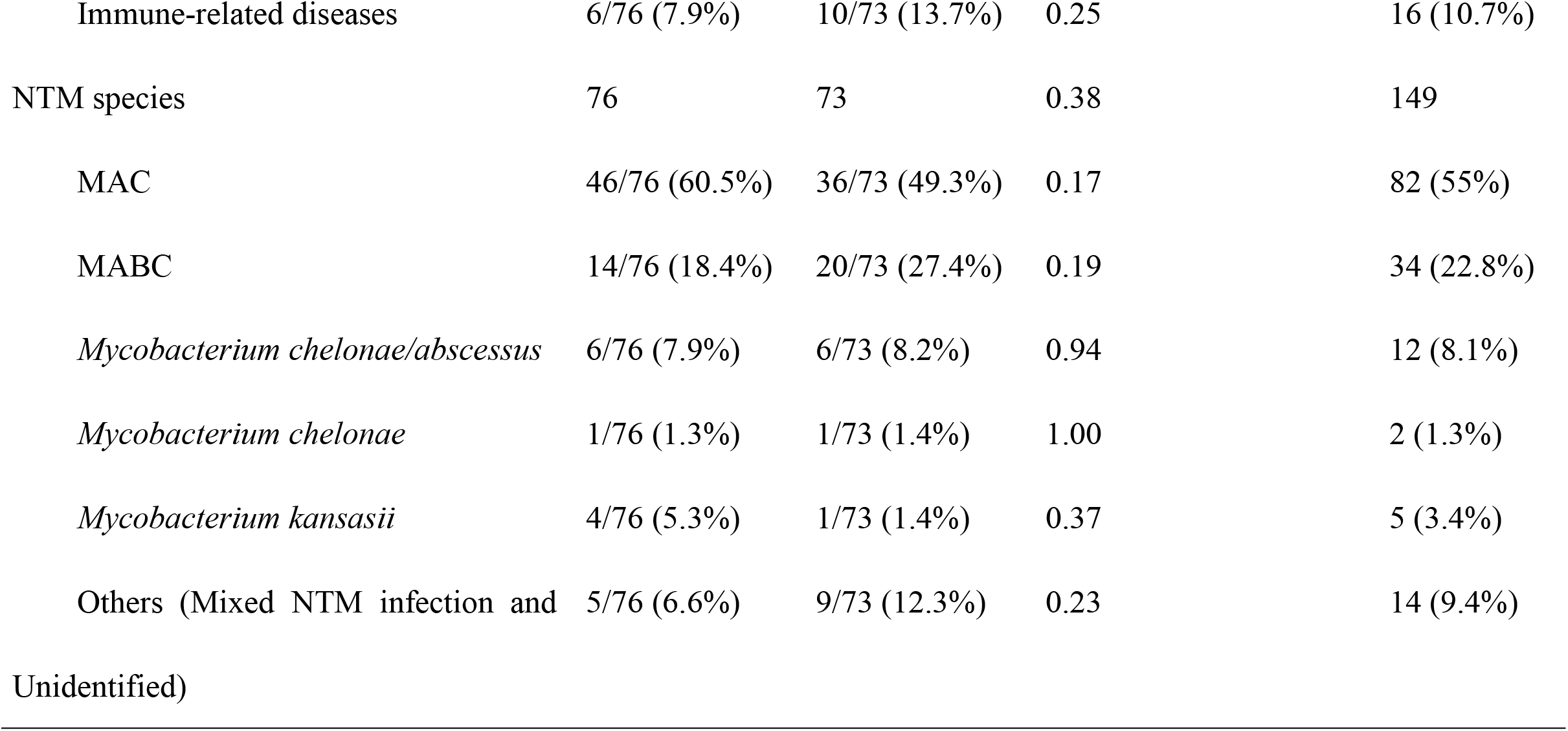
Patient characteristics in control group and sitafloxacin group.

The distribution of NTM species was also comparable between the two groups (*P* = 0.38). The most common species was MAC (60.5% vs. 49.3%, *P* = 0.17), followed by MABC (18.4% vs. 27.4%, *P* = 0.19), *M. chelonae/abscessus* (7.9% vs. 8.2%, *P* = 0.94), *M. chelonae* (1.3% vs. 1.4%, *P* = 1.00), *M. kansasii* (5.3% vs. 1.4%, *P* = 0.37), and other mixed or unidentified NTM infections (6.6% vs. 12.3%, *P* = 0.23).

### Efficacy and safety outcomes

In terms of microbiological efficacy (**Table 2**), the median time to conversion was significantly shorter in the sitafloxacin group than the control group (195 days [95% CI: 146.8-243.2] vs. 292 days [95% CI: 233.4-350.6], *P* < 0.001). In the subgroup analysis, among patients with MAC infection, the median time to culture conversion was significantly shorter in the sitafloxacin group compared to the control group (195 days [95% CI: 114.4-275.6] vs. 288 days [95% CI: 223.7-352.3], *P* < 0.001). In contrast, among patients with MABC infection, the median time to conversion did not differ significantly between the two groups (328 days [95% CI: 138.7-517.3] vs. 299 days [95% CI: 90.5-507.5], *P* = 0.54).

**Table 2.** The efficacy and safety of control group 524 and sitafloxacin group.

| Outcome | Control, n (%) | Sitafloxacin, n (%) | P Value |
| --- | --- | --- | --- |
| **Efficacy** |  |  |  |
| Microbiological examinations |  |  |  |
| Time of conversion [median (95% CI), day] | 292 (233.4-350.6) | 195 (146.8-243.2) | < 0.001 |
| MAC | 288 (223.7-352.3) | 195 (114.4-275.6) | < 0.001 |
| MABC | 299 (90.5-507.5) | 328 (138.7-517.3) | 0.54 |
| Conversion post-therapy | 15/68 (22.1%) | 28/52 (53.8%) | < 0.001 |
| MAC | 9/42 (21.4%) | 15/27 (55.6%) | 0.004 |
| MABC | 3/14 (21.4%) | 6/16 (37.5%) | 0.44 |
| Sputum smear positivity | 38/84 (45.2%) | 12/45 (26.7%) | 0.04 |
| Sputum/BALF culture positivity | 86/263 (32.7%) | 51/227 (22.5%) | 0.01 |
| DNA of mycobacterium | 53/77 (68.8%) | 58/85 (68.2%) | 0.94 |
| NGS of sputum/BALF | 38/40 (95%) | 68/71 (95.8%) | 1.00 |
| Xpert to detect *M. tuberculosis* | 0/79 | 0/75 | 0.93 |
| Completion of treatment |  |  |  |
| Time [median (95% CI), day] | 809 (338.0-1280.0) | 406 (371.0-441.0) | < 0.001 |
| Rate | 26/76 (34.2%) | 25/73 (34.2%) | 1.00 |
| Radiological improvement | 22/61 (36.1%) | 30/55 (54.5%) | 0.046 |
| **Safety** |  |  |  |
| Adverse events |  |  |  |
| Gastrointestinal reactions | 11/76 (14.5%) | 11/73 (15.1%) | 0.92 |
| Hepatic or renal dysfunction | 25/76 (32.9%) | 25/73 (34.2%) | 0.86 |
| Hematologic abnormalities | 11/76 (14.5%) | 18/73 (24.7%) | 0.12 |
| Arthralgia | 2/76 (2.6%) | 4/73 (5.5%) | 0.44 |
| Rash | 3/76 (3.9%) | 12/73 (16.4%) | 0.01 |

At the median culture conversion time point of 195 days from the initiation of sitafloxacin, the sitafloxacin group demonstrated a significantly higher sputum/BALF conversion rate compared to the control group at the corresponding 195-day mark from the initiation of NTM treatment (53.8% [28/52] vs. 22.1% [15/68], *P* < 0.001). The subgroup analysis based on NTM species revealed that, among patients with MAC infection, the sitafloxacin group demonstrated a significantly higher post-therapy conversion rate compared to the control group (55.6% [15/27] vs. 21.4% [9/42], *P* = 0.004). For MABC patients, while the sitafloxacin group had a higher conversion rate compared with the control group (37.5% [6/16] vs. 21.4% [3/14]) it did not reach statistically significant difference (*P* = 0.44).

The proportion of patients with positive sputum smears (26.7% [38/84] vs. 45.2% [12/45], *P* = 0.04) and cultures from sputum or BALF samples (22.5% [51/227] vs. 32.7% [86/263], *P* = 0.01) was significantly lower in the sitafloxacin group than the control group (**Table 2**). Regarding treatment completion, the median duration to treatment completion was significantly shorter in the sitafloxacin group (406 days [95% CI: 371.0-441.0] vs. 809 days [95% CI: 338.0-1280.0], *P* < 0.001), but the rate of treatment completion was not statistically significant between two groups (34.2% [25/73] vs. 34.2% [26/76], *P* = 1.00). Other molecular diagnostic results, including detection of *M. tuberculosis* DNA, next-generation sequencing (NGS), and Xpert MTB/RIF testing, showed no significant differences between the two groups (*P* > 0.05).

Patients in the sitafloxacin group demonstrated a significantly higher rate of radiological improvement compared to the control group (54.5% [30/55] vs. 36.1% [22/61], *P* = 0.046) (**Table 2**), as assessed by pre- and post-treatment CT scans. To visually illustrate the clinical efficacy of sitafloxacin, **Figure 2** presents a representative case. The patient initially received a non-sitafloxacin regimen, and baseline CT revealed extensive pulmonary lesions (**Figure 2A**). After 12 months of conventional therapy which did not include sitafloxacin (**Figure 2B**), the patient exhibited radiological worsening, consistent with treatment failure. Consequently, the regimen was switched to include sitafloxacin. Remarkable radiological improvement was noted after 7 months of sitafloxacin-based therapy (**Figure 2C**). This case further underscores the potential efficacy of sitafloxacin in patients with suboptimal response to standard regimens.

**Figure 2.**
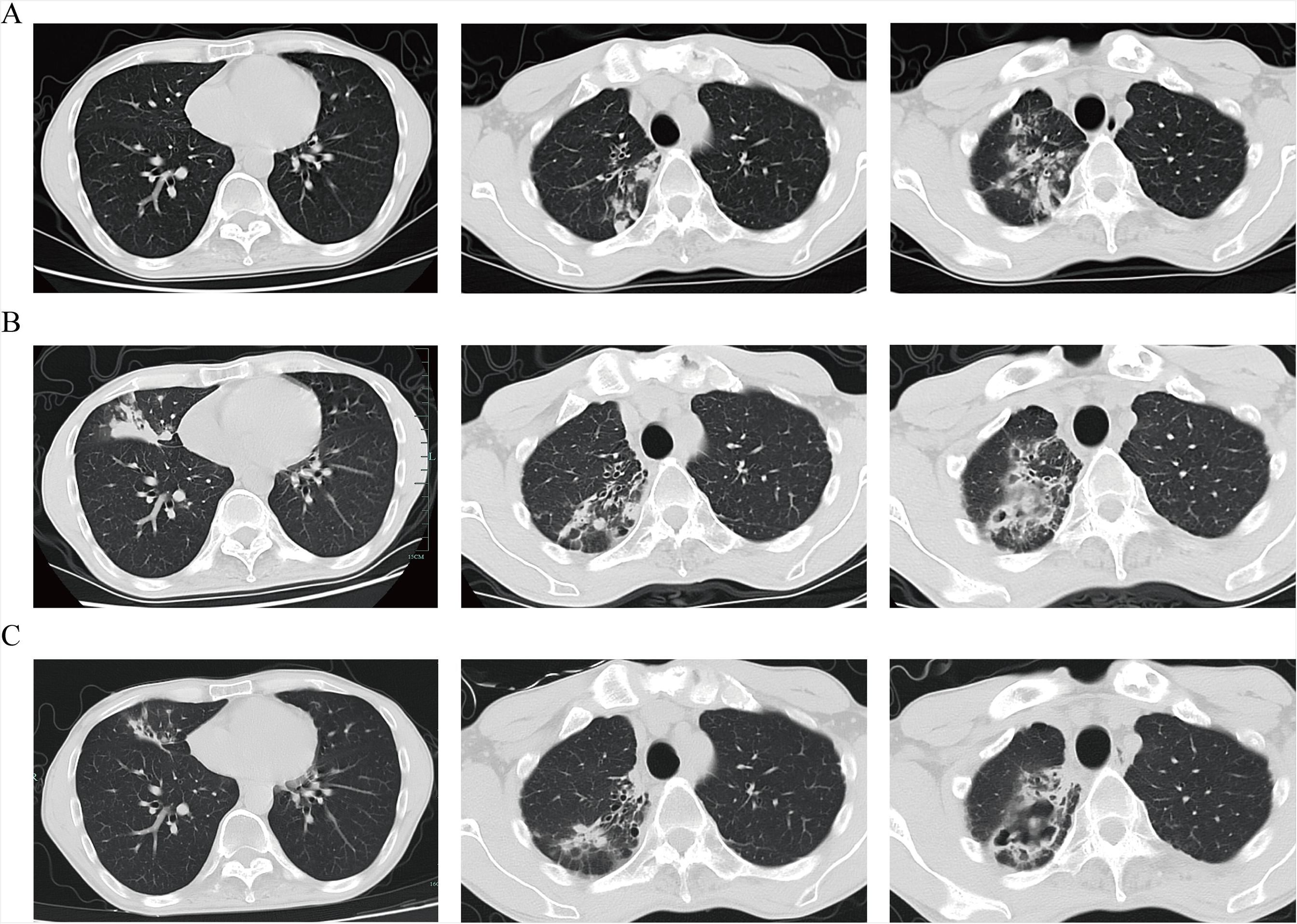
Representative CT images of a patient from the sitafloxacin group who switched regimens due to treatment failure. (A) Axial CT scans at three representative levels before initiation of NTM therapy. (B) Corresponding CT images after 12 months of treatment with a non-sitafloxacin-containing regimen, demonstrating radiological progression. (C) CT images at the same three levels following 7 months of treatment with a sitafloxacin-containing regimen, showing substantial improvement. This case illustrates radiological benefit after switching to sitafloxacin due to prior regimen failure.

Adverse events were generally comparable between the sitafloxacin and control groups (**Table 2**), with no statistically significant differences observed in most categories. Gastrointestinal reactions occurred in 14.5% (11/76) of patients in the control group and 15.1% (11/73) in the sitafloxacin group (*P* = 0.92). Hepatic or renal dysfunction was noted in 32.9% (25/76) and 34.2% (25/73) of patients, respectively (*P* = 0.86). Hematologic abnormalities were slightly more frequent in the sitafloxacin group (24.7%, 18/73) compared to the control group (14.5%, 11/76), although the difference did not reach statistical significance ((*P* = 0.12). Arthralgia was reported in 2.6% (2/76) of control patients and 5.5% (4/73) of patients in the sitafloxacin group (*P* = 0.44). Notably, rash occurred significantly more frequently in the sitafloxacin group (16.4%, 12/73) than in the control group (3.9%, 3/76), with a statistically significant difference (*P* = 0.01).

### Treatment profile of the sitafloxacin cohort

Among the 73 patients in the sitafloxacin group, the most common indication for sitafloxacin initiation was treatment failure of a prior regimen (n = 41, 56.2%), in which the conversion with prior-sitafloxacin therapy was only 4.9%, while in contrast, the conversion rate post-sitafloxacin therapy was 53.7%, calculated over the entire follow-up period rather than at the previously described median conversion time point of 195 days (**Table 3**). When sitafloxacin was used as the initial first-line therapy in 15 patients (20.5%) the conversion rate was very high at 86.7%. However, 17 patients (23.3%) who received sitafloxacin due to discontinuation of previous regimens resulting from ADRs had post-sitafloxacin therapy conversion rate of 52.9%, and the most frequent AE was gastrointestinal reactions (3/9). Regarding dosage, the vast majority of patients (n = 69, 94.5%) received 100 mg per day. Only 3 patients (4.1%) were treated with 200 mg daily, and 1 patient (1.4%) received a 50 mg daily dose. As for treatment discontinuation, 24 patients (32.9%) completed the sitafloxacin therapy, while 12 patients (16.4%) discontinued due to adverse events (four due to rash, two arthralgia, two fatigue, two gastrointestinal reactions, one hepatic dysfunction and one fungal infection).

**Table 3.**
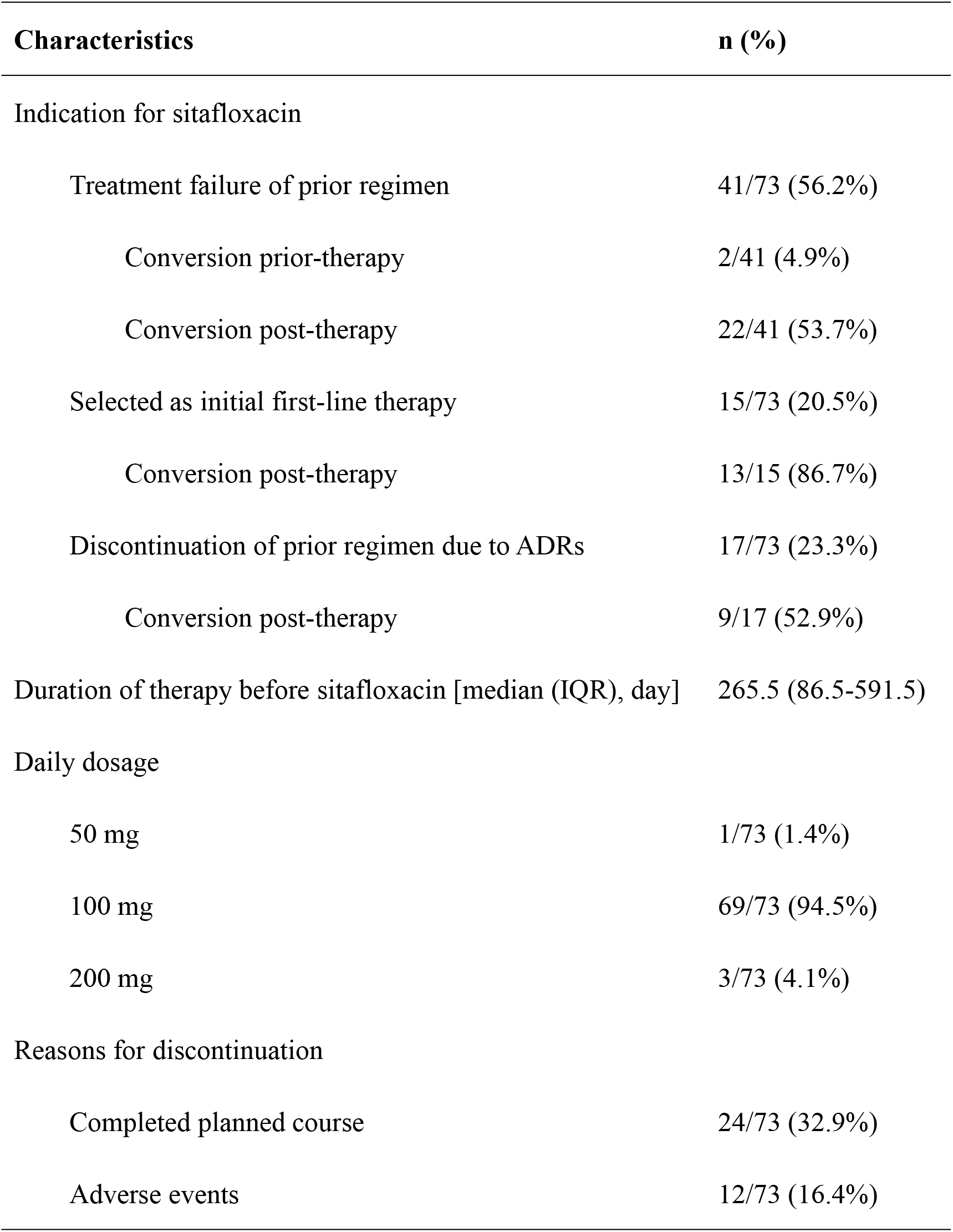
Characteristics of sitafloxacin administration in NTM patients.

| Characteristics | n (%) |
| --- | --- |
| Indication for sitafloxacin |  |
| Treatment failure of prior regimen | 41/73 (56.2%) |
| Conversion prior-therapy | 2/41 (4.9%) |
| Conversion post-therapy | 22/41 (53.7%) |
| Selected as initial first-line therapy | 15/73 (20.5%) |
| Conversion post-therapy | 13/15 (86.7%) |
| Discontinuation of prior regimen due to ADRs | 17/73 (23.3%) |
| Conversion post-therapy | 9/17 (52.9%) |
| Duration of therapy before sitafloxacin [median (IQR), day] | 265.5 (86.5-591.5) |
| Daily dosage |  |
| 50 mg | 1/73 (1.4%) |
| 100 mg | 69/73 (94.5%) |
| 200 mg | 3/73 (4.1%) |
| Reasons for discontinuation |  |
| Completed planned course | 24/73 (32.9%) |
| Adverse events | 12/73 (16.4%) |

### Antibiotic treatment regimens

To comprehensively evaluate treatment strategies, we compared the frequencies of antibiotic use between the control and sitafloxacin groups following sitafloxacin administration (**Table S1**). The sitafloxacin group exhibited a statistically significant reduction in the use of moxifloxacin, levofloxacin, rifamycins (including rifampin, rifabutin, and rifapentine), and ethambutol compared to the control group (adjusted *P* ≤ 0.01). Although azithromycin and amikacin usage declined in the sitafloxacin group, these differences did not retain significance after Bonferroni correction (adjusted *P* = 0.06 and 0.58, respectively). The frequency of linezolid use remained comparable between groups, while slight non-significant increases were noted for tetracyclines, clofazimine, and bedaquiline in the sitafloxacin group relative to controls. These results delineate a distinctive antibiotic usage profile associated with sitafloxacin treatment, characterized primarily by decreased reliance on select macrolides, rifamycins, and ethambutol.

## Discussion

In this multi-center retrospective cohort treatment study of NTM pulmonary disease, sitafloxacin-containing regimens achieved 53.8% sputum/BALF culture conversion, significantly surpassing outcomes with the current guideline-based therapies (22.1% conversion rates in matched cohorts). Furthermore, the median time to culture conversion was reduced by 97 days in the sitafloxacin treatment group compared to the guideline-based therapy (195 vs. 292 days), with significant radiographic improvement in the sitafloxacin group (*P* ˂ 0.05). These benefits were attained without excess drug toxicity.

Standard macrolide-based multidrug regimens for MAC pulmonary disease achieve sputum culture conversion rates of 67-84% in macrolide-susceptible patients but only 5-36% in macrolide-resistant cases, with median conversion times typically exceeding 12 months.^7^ In contrast, the present study demonstrated a notably shorter median time to conversion (195 vs. 288 days, *P* ˂ 0.001) and a higher overall conversion rate (55.6% vs. 21.4%, *P* = 0.004) in the sitafloxacin treatment group. These findings suggest that the incorporation of sitafloxacin may enhance therapeutic efficacy and accelerate microbiological clearance in patients with MAC lung disease, particularly in treatment-refractory cases. Our findings align with a previous study demonstrating the potential efficacy of sitafloxacin in treating refractory MAC lung disease. For instance, Asakura et al.^18^ reported a 23% sputum culture conversion rate and 19% radiological improvement in 31 patients with refractory MAC-LD treated with an sitafloxacin-containing regimen, highlighting its role as a salvage therapy in difficult-to-treat cases. Similarly, our findings further support the clinical benefit of sitafloxacin in the management of NTM pulmonary disease. Consistent with previous findings, our study reported mild AEs, underscoring sitafloxacin’s favorable safety profile in these clinical scenarios.^18^

However, our study offers several advantages. In contrast to the study conducted by Asakura et al.,^18^ our study incorporated a control group without sitafloxacin exposure and also included a larger sample size (73 vs. 31). In addition, this investigation provides novel evidence regarding the treatment duration dynamics of sitafloxacin therapy compared to the previous study.^18^ The significantly shorter median treatment time observed in the sitafloxacin group (406 days) compared with the control group (809 days) suggests a potential disease-modifying effect that accelerated the treatment time in NTM patients. Furthermore, radiographic improvement served as an early-response indicator. The significantly higher radiological response rate with sitafloxacin (54.5% vs. 36.1%; *P* = 0.046), particularly the accelerated fibrosis resolution observed in Figure 2, provides objective imaging evidence for its improved bactericidal activity. Moreover, our analysis stratified treatment indications and corresponding conversion rates, revealing that patients who received sitafloxacin due to prior treatment failure still achieved a high post-therapy conversion rate (56.2%), whereas earlier studies did not evaluate sitafloxacin efficacy according to treatment background or conversion status.^18,20^

In our study, sitafloxacin showed a significant microbiological benefit in patients with MAC infections, consistent with previous in vitro studies reporting enhanced activity of sitafloxacin against MAC isolates.^20^ However, for MABC infections, while the sitafloxacin group demonstrated a higher culture conversion rate than the control group (37.5% vs. 21.4%), the difference did not reach statistical significance (*P* = 0.44). Several factors may account for this differential response for MAC versus MABC infections. Firstly, MABC is known to exhibit stronger efflux pump activity,^21,22^ which contributes to lower intracellular drug accumulation and may limit the efficacy of fluoroquinolones, including sitafloxacin. Secondly, the duration of sitafloxacin therapy (3 months or more) in our MABC cohort may have been inadequate for achieving sputum conversion, considering that MABC infections are more difficult to treat and require prolonged treatment due to their intrinsic resistance and stronger efflux activity. Thirdly, the finding of a statistically insignificant difference may be influenced by the limited smaller cohort size (16 vs. 14) in MABC patients and possible selection bias, which warrants cautious interpretation and further investigation. Future prospective studies with longer sitafloxacin treatment regimens and more patients are needed to address the potential benefit of this drug for the treatment of MABC infections.

Sitafloxacin’s current approved indications are limited to infections caused by *Pseudomonas aeruginosa*, refractory urinary tract infections, and community-acquired pneumonia. Its use in NTM pulmonary disease remains investigational, and clinical trials for tuberculosis are still ongoing. In this study, the enhanced efficacy observed in the sitafloxacin group is likely attributable to the unique antimicrobial property of sitafloxacin, rather than the contribution of other background antibiotics. The treatment regimens used in both groups were largely comparable, and no other specific agent was disproportionately represented in the sitafloxacin group (Table S1). In fact, several antibiotics such as rifamycins and ethambutol and other fluoroquinolones were less frequently used in the sitafloxacin group (Table S1), ruling out the possibility that the improved efficacy of the sitafloxacin group is due to background drugs.

Several mechanisms may explain sitafloxacin’s superior activity. First, sitafloxacin exhibits significantly lower MIC against *M. abscessus* and *M. avium* (both 0.5 mg/L) than moxifloxacin (4 mg/L and 1 mg/L, respectively), indicating enhanced potency at lower tissue concentrations.^20^ Second, the therapeutic management of NTM infections is notably compromised by their recalcitrant nature, particularly given the extreme drug tolerance exhibited by *M. abscessus* persisters.^23^ This inherent resistance phenotype substantially diminishes treatment efficacy.^24,25^ Crucially, our preliminary investigations indicate sitafloxacin has unique capacity to eradicate drug tolerant persisters (Y Zhang, unpublished data), suggesting a mechanistic breakthrough against persistent organisms. Third, unlike older quinolones that target to DNA gyrase or topoisomerase IV, the dual targeting mechanism of sitafloxacin not only improves its antimicrobial activity but also delays the development of resistance,^26–28^ Therefore, these properties support the proposal that sitafloxacin plays a primary role in the improved outcomes observed in our study, independent of other concurrent antibiotics.

Admittedly, our findings must be interpreted in the context of several methodological limitations inherent to retrospective studies. First, although all patients in the sitafloxacin group received the drug for at least three months, the actual treatment duration varied between individuals. Since prolonged therapy may lead to better clinical outcomes, those receiving sitafloxacin for longer periods might have derived greater benefit. This heterogeneity could potentially bias the overall efficacy assessment. Future prospective studies should standardize treatment durations to better evaluate the true therapeutic potential of sitafloxacin. Second, the interpretation of treatment completion rates is inherently limited by the predefined observational time frame. At the time of data cutoff, only a portion of patients (34.2% in both groups) had reached the therapeutic endpoint. This time-restricted evaluation may introduce selection bias and render the current comparison of completion rates provisional. Longer follow-up periods are necessary to reassess treatment completion dynamics once more patients reach the intended endpoint. Third, variations in background antibiotic regimens between groups may have confounded the observed treatment outcomes. In real-world practice, patients with NTM pulmonary disease often receive multiple concomitant antibiotics. Differences in baseline regimens and subsequent adjustments could influence therapeutic responses, making it difficult to isolate the independent effect of sitafloxacin. Due to the inability to strictly control background treatments in this study, caution is warranted when attributing observed benefits solely to sitafloxacin. Future research should aim to standardize concomitant treatments or employ randomized controlled trial designs to minimize confounding. In summary, our findings highlight the urgent need for large-scale, prospective randomized controlled trials comparing sitafloxacin-containing regimens with current guideline-recommended therapies. Such studies are essential to establish robust evidence for the optimal clinical use of sitafloxacin in the management of NTM pulmonary disease.

## Conclusions

This retrospective cohort study demonstrates that incorporating sitafloxacin into treatment regimens significantly accelerates sputum/BALF culture conversion rate and time, as well as enhances radiographic improvement in patients with NTM pulmonary disease, without increasing severe AEs, underscoring its potential as a first-line alternative for more effective treatment of NTM infections.

## Data Availability

All data produced in the present work are contained in the manuscript

## Author Contributions

Yiru Zhang, Hongxin Fu and Kefan Bi contributed to the research conception and design. Yiru Zhang, Hongxin Fu, Chenxia Zhang, Bihan Xu, Meifang Yang, Haiying Yu, Yongtao Li, Jing Guo and Wei Wu collect the data. Yiru Zhang and Hongxin Fu performed data analysis and wrote the manuscript. Ying Zhang and Kaijin Xu contributed to the review and editing. All the authors have given approval to the final version of the manuscript.

## Ethics Approval and Consent to Participate

This study is approved by the Ethics Committee of the First Affiliated Hospital, Zhejiang University School of Medicine (Approval number.: 2025B-IIT Ethics Approval No. 0666).

## Availability of Data and Material

All data generated or analyzed during this study are included in this article.

