## Supplemental Table 1 for "Efficacy and Safety of Sitafloxacin-Containing Regimen for Improved Treatment of Nontuberculous Mycobacterial Lung Disease: A Multi-Center Retrospective Study"

[1] State Key Laboratory for Diagnosis and Treatment of Infectious Diseases, National Clinical Research Center for Infectious Diseases, China-Singapore Belt and Road Joint Laboratory on Infection Research and Drug Development, National Medical Center for Infectious Diseases, Collaborative Innovation Center for Diagnosis and Treatment of Infectious Diseases, The First Affiliated Hospital, Zhejiang University School of Medicine, Hangzhou, Zhejiang 310003, China

[2] Yuhang Institute for Collaborative Innovation and Translational Research in Life Sciences and Technology

[3] Jinan Microecological Biomedicine Shandong Laboratory, Jinan 250117, China

[#] Yiru Zhang and Hongxin Fu contributed equally to this work

*Corresponding authors:

**Table S1** Summary of antibiotic classes and usage frequency in the control and sitafloxacin groups (after sitafloxacin administration)

| **Antibiotic** | **Control (n=73)** | **Sitafloxacin (n=76)** | ***P* Value** | **Adjusted *P*** |
|---|---|---|---|---|
| Azithromycin | 61 (80.3%) | 48 (65.8%) | 0.005 | 0.06 |
| Clarithromycin | 26 (34.2%) | 18 (24.7%) | 0.11 | 1.00 |
| Moxifloxacin | 54 (71.1%) | 1 (1.4%) | < 0.001 | <0.01 |
| Levofloxacin | 31 (40.8%) | 0 | < 0.001 | <0.01 |
| Amikacin | 26 (34.2%) | 16 (21.9%) | 0.048 | 0.58 |
| Linezolid | 35 (46.1%) | 34 (46.6%) | 0.70 | 1.00 |
| Rifamycins | 60 (78.9%) | 44 (57.9%) | 0.001 | 0.01 |
| Ethambutol | 35 (46.1%) | 10 (13.7%) | < 0.001 | <0.01 |
| Carbapenems | 23 (30.3%) | 17 (23.3%) | 0.21 | 1.00 |
| Tetracyclines | 13 (17.1%) | 19 (26%) | 0.29 | 1.00 |
| Clofazimine | 2 (2.6%) | 4 (5.5%) | 0.68 | 1.00 |
| Bedaquiline | 1 (1.3%) | 5 (6.8%) | 0.21 | 1.00 |

Note: Adjusted *P* values were calculated using the Bonferroni correction, with a significance threshold set at $\alpha = 0.0042$.
